# Proteomic signatures and machine learning based-prediction models for cardiovascular risk in survivors of myocardial infarction

**DOI:** 10.1101/2025.10.20.25338422

**Authors:** Shizhen Xiang, Yuge Ye, Xi Cao, Huidan Zeng, Yunlong Guan, Siyu Zhu, Xiangjing Liu, Da Luo, Yifan Kong, Zhonghe Shao, Bofang Zhang, Xingjie Hao

**Author notes:** Correspondence should be addressed to Xingjie Hao < > or Bofang Zhang < >. Shizhen Xiang and Yuge Ye contributed equally.

## Abstract

**Background:** Survivors of myocardial infarction (MI) are still at risk for adverse long-term outcomes such as all-cause mortality, heart failure (HF), and ischemic stroke (IS) after acute phase treatment.

**Aims:** This study aimed to identify specific protein markers and construct risk prediction models for the main cardiovascular events in survivors of MI.

**Methods:** A total of 30,135 survivors of MI were included in this study, all of whom had available follow-up data from the UK Biobank (UKB). Multivariate Cox regression analysis was used to assess the clinical associations between plasma proteins and MI-related outcomes, including all-cause mortality, HF and IS. Subsequently, prediction models with machine learning were constructed based on the plasma protein levels to further evaluate these associations.

**Results:** We identified 570 proteins significantly associated with all-cause mortality, 172 with HF, and 13 with IS in survivors of MI. Among these proteins, 12 proteins were associated with three outcomes (*P* < 1.71◊10^-5^). Pathway enrichment analysis showed that these proteins were mainly involved in pathophysiological processes such as inflammatory response, fibrosis and myocardial remodeling. Machine learning models based on 117, 73 and 82 plasma protein showed good predictive performance for all-cause mortality (XGBoost: AUC = 0.79), HF (LightGBM: AUC = 0.81) and IS (Random Forest: AUC = 0.76) in survivors of MI, respectively. Finally, we systematically identified 52 plasma proteins associated with all-cause mortality, 14 with HF, and 4 with IS in survivors of MI through integrated Cox regression and machine learning modeling.

**Conclusion:** Our integrated study with predictive modeling have identified the plasma protein biomarkers associated with adverse outcomes in survivors of MI, and subsequently developed predictive models to facilitate early risk stratification.

## Introduction

Myocardial infarction (MI), the most severe clinical manifestation of coronary artery disease (CAD)^[1]^, remains the leading cause of sudden cardiac death (SCD) worldwide^[2]^. Although significant improvements in acute-phase survival through thrombolysis and interventional therapy, MI survivors still has a major challenge for long-term prognosis^[3]^. Actually, MI survivors remain at high risk for adverse outcomes including all-cause mortality, heart failure (HF), and ischemic stroke (IS), with these complications severely impacting their quality of life^[4–6]^.

Traditional biomarkers, such as NT-proBNP, which reflects myocardial wall stress, and CRP, an indicator of systemic inflammation, are commonly used to predict adverse outcomes like HF and death in survivors of MI. However, these markers alone cannot fully explain the marked heterogeneity in long-term outcomes among survivors of MI. Although several emerging protein biomarkers, such as GDF-15 and IL-6^[7,8]^, have shown promise in cardiovascular risk prediction, current evidence is constrained by small sample sizes, insufficient adjustment for confounders, and a predominant focus on single outcomes^[6,9]^. Systematic and large-scale investigations specifically targeting MI survivors remain scarce. In parallel, while machine learning techniques hold great potential for capturing complex patterns in proteomic data, their application in MI-related prognostic research remains in the early stages, and a robust analytical framework has yet to be established.

In this context, identifying novel biomarkers and developing refined predictive tools tailored to MI survivors has become increasingly important. Proteomics offers a promising solution, as it captures dynamic biological processes that directly reflect underlying pathophysiological changes, including myocardial remodeling and chronic inflammation^[9^ ^10^]. With the advent of high-throughput platforms, such as SomaScan and Olink, large-scale screening of plasma proteins provided new possibilities for predicting adverse outcomes in survivors of MI.

Here, we investigated the associations between plasma proteins and multiple clinical outcomes, including all-cause mortality, HF, and IS in survivors of MI using data from the UK Biobank (UKB). This study aimed to identify prognostic protein biomarkers, explore their underlying biological pathways, and develop predictive models tailored to the survivors of MI, with the ultimate goal of improving long-term prognosis and enhancing quality of life for survivors of MI.

## Methods

### Study population and overall design

Data for this study were obtained from the UKB, a large-scale, NHS-supported prospective cohort that recruited more than 500,000 participants aged between 40 and 69 years across the United Kingdom between 2006 and 2010, with ongoing long-term follow-up. All participants provided written informed consent, and the study was approved by the North West Multi-Center Research Ethics Committee (Research Ethics Committee reference: 11/NW/0382). Participant selection and the overall analytical framework are presented in Figure 1. The study population consisted of 11,851 MI survivors. These participants were identified prior to the baseline assessment using the algorithm defined by the UKB (Field ID: 42000). The algorithmically-defined outcome measures include data on probable cases of selected health conditions, which are derived from algorithmic combinations of coded information. Specifically, this includes baseline assessment data from the UKB (encompassing participants’ self-reported medical conditions, surgeries, and medication use), as well as linked hospital admission data (diagnostic and procedural information) and death registry data. To ensure that HF and IS cases were incident events, the diagnosis date was required to occur strictly after the baseline date. Thus, participants with a prior recorded diagnosis of HF or IS before baseline were excluded from the respective outcome analyses. All-cause mortality included all deaths occurring during the follow-up period. The application number of UKB in our study was 88159.

**Figure 1.**
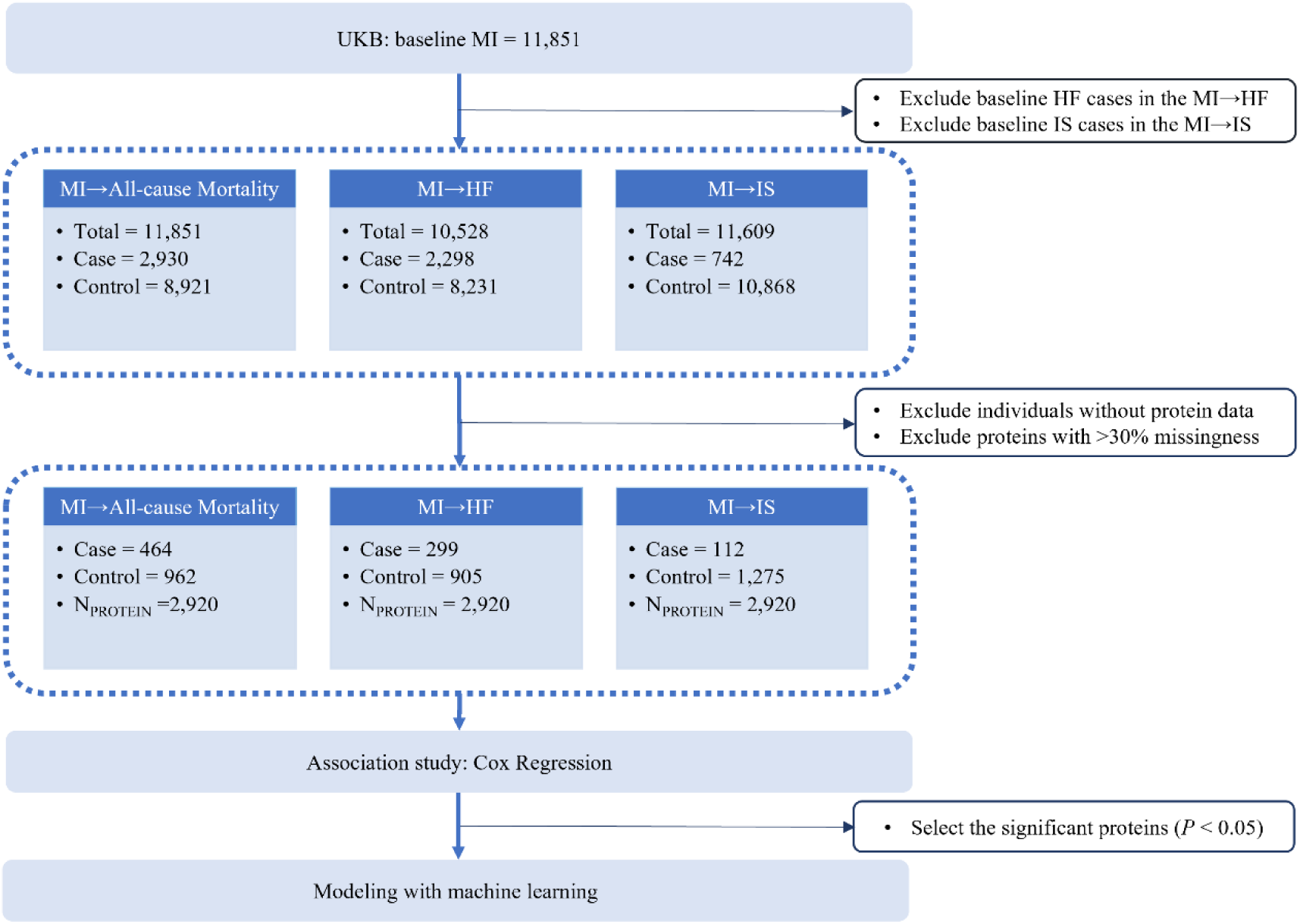
Flowchart of the overall study design. 30,135 survivors of MI from UK Biobank were included, excluding those with prevalent HF or IS in MI→HF and MI→IS cohorts. Remaining patients were stratified into MI→ all-cause mortality, MI→HF, and MI→IS cohorts. Proteins with > 30% missing values were removed. Final cohorts underwent observational analysis and machine learning. MI: Myocardial Infarction; HF: Heart Failure; IS: Ischemic Stroke.

### Measurement of exposures

In the UKB, plasma protein levels were measured for blood samples collected at baseline during initial recruitment (2006∼2010). High-throughput proteomic profiling was conducted by Olink using a proximity extension assay, which enables the simultaneous quantification of a wide range of proteins with high specificity and sensitivity. Exposure variables in this study were based on the concentrations of 2,923 plasma proteins provided by the UKB Plasma Proteome Project (UKB-PPP).

Proteins with more than 30% missing values were excluded, resulting in a final dataset of 2,920 proteins. For the remaining protein data, missing values were imputed using the k-nearest neighbors (KNN) algorithm, implemented via the *knnImputation* function in R. The number of neighbors (K) was set to 3 to optimize the use of sample similarity in imputing missing values and to minimize potential bias introduced by missing data.

### Ascertainment of outcomes

The primary outcomes of this study were all-cause mortality, new-onset HF, and new-onset IS. Outcome events were ascertained through linkage to the UK Biobank’s comprehensive electronic health record system, which includes primary care data, hospital inpatient records, and national death registry data. The corresponding Field-ID codes and ICD-10 codes used to define each outcome are provided in the Supplementary Data 1-2. Three sub-cohorts were constructed: the MI→all-cause mortality cohort included 464 deaths and 962 controls (n = 1,426), the MI→HF cohort included 299 HF events and 905 controls (n = 1,204), and the MI→IS cohort included 112 IS events and 1,275 controls (n = 1,387). Follow-up time was calculated as the minimum of the time to the outcome event, time to censoring, or time from baseline to the end of follow-up, and was expressed in years.

### Assessment of covariates

Baseline characteristics of the study population included demographic variables (age, sex), socioeconomic factors (Townsend deprivation index, education level), and lifestyle factors (body mass index [BMI], smoking status, alcohol consumption, and physical activity). For detailed information, please refer to Table 1. For continuous variables, missing values were imputed using the median. For categorical variables, missing values were treated as a separate category labeled “Missing”.

**Table 1:**
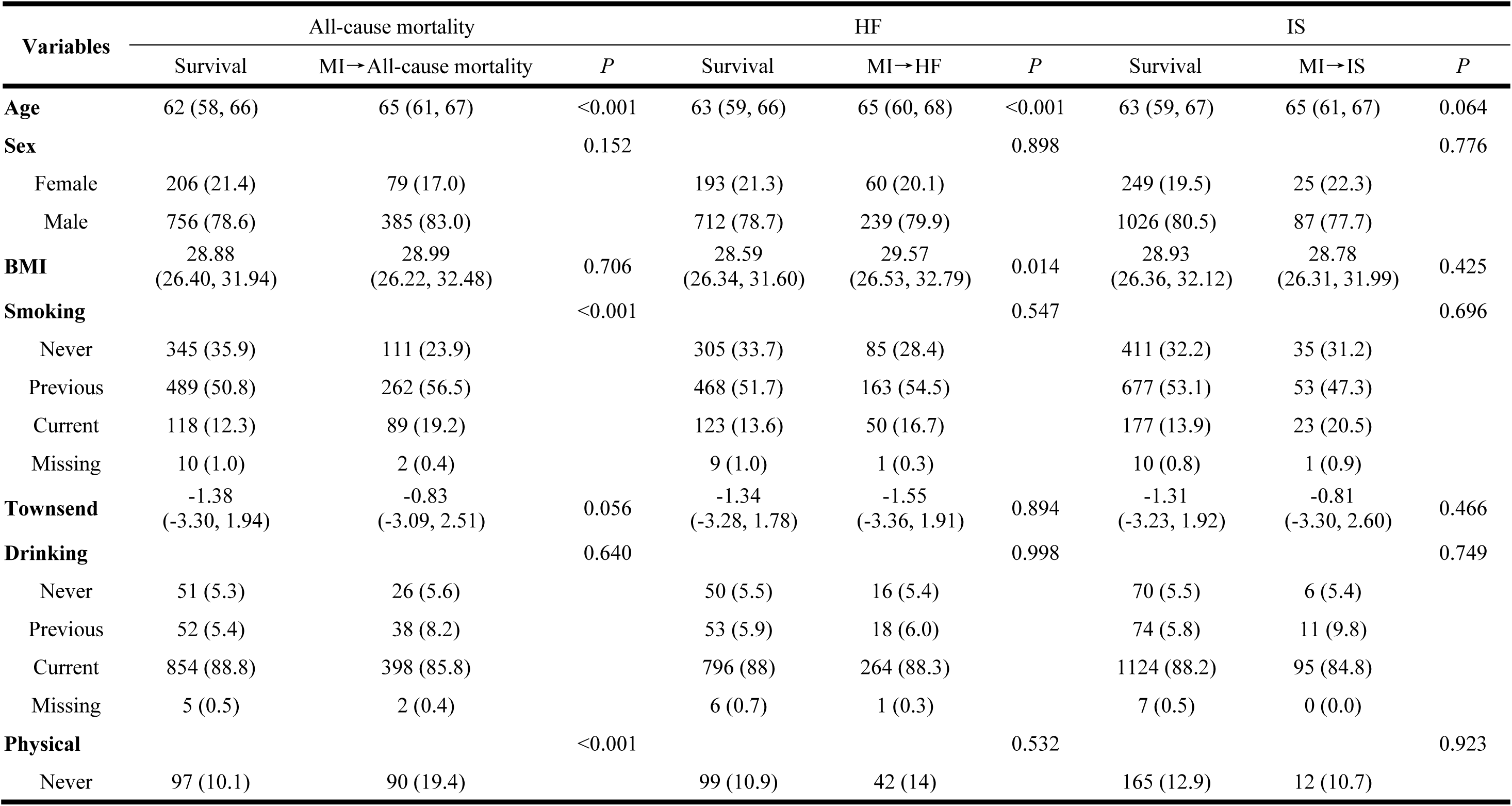
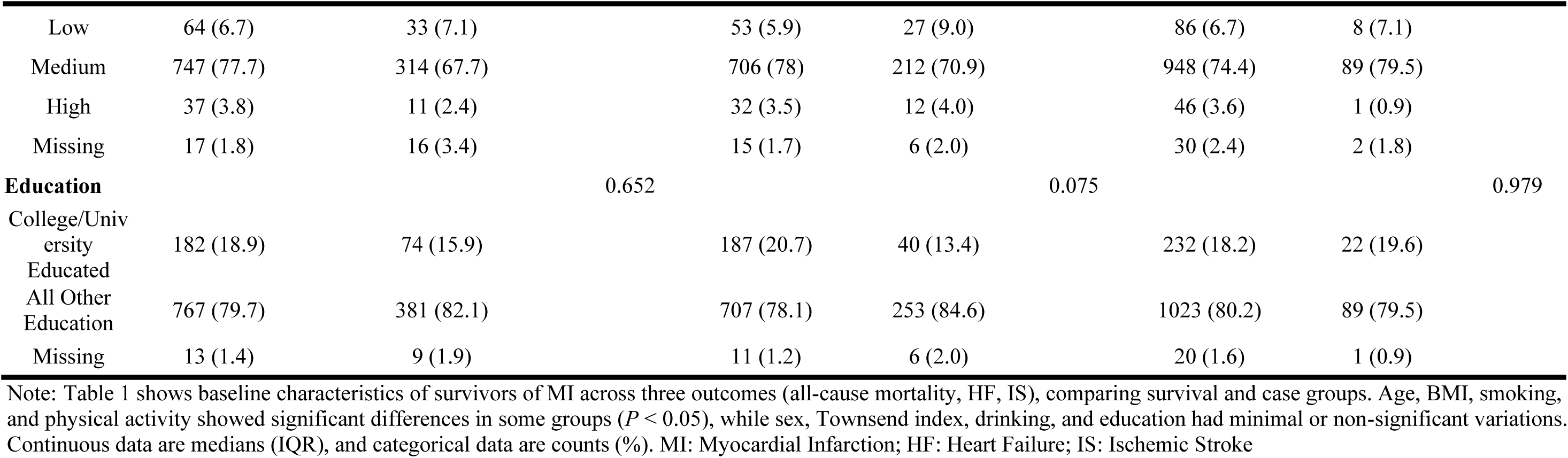
Baseline characteristics.

### Statistical analysis

The baseline characteristics of participants were described based on MI-related outcome status. Continuous variables are presented as median (interquartile range, IQR) and categorical variables as frequency (percentage). Categorical variables were compared using the Chi-square test, while continuous variables were compared using the Wilcoxon rank-sum test.

Plasma protein levels were first inverse-normal transformed. Multivariable Cox proportional hazards models were then used to estimate hazard ratios (HRs) and 95% confidence intervals (CIs) for the three outcomes. The models were adjusted for potential confounders, including age, sex, body mass index (BMI), smoking status, alcohol consumption, education level, Townsend deprivation index, and physical activity.

To ensure the robustness of the findings, several sensitivity analyses were conducted. First, nonlinear associations between plasma protein levels and outcomes were assessed using restricted cubic splines with three knots, adjusting for the same covariates as in the multivariable Cox model. Next, subgroup analyses stratified by age and sex were performed to evaluate the consistency of associations across different demographic groups. Finally, to account for competing risks of death, Fine-Gray subdistribution hazard models were applied in the MI→HF and MI→IS cohorts.

### Pathway enrichment analysis

Pathway enrichment analysis was conducted for proteins significantly associated with outcomes, covering biological process (BP), molecular function (MF), and cellular component (CC) categories. These analyses used all 2,923 UKB-PPP proteins studied here as the background gene set.

### Risk prediction modeling with machine learning strategy

In this study, a two-stage modeling strategy was used to systematically screen and evaluate predictive protein markers. Protein markers significantly associated with endpoint events were first initially screened by the above univariate Cox regression analysis (*P* < 0.05), and KNN interpolation was performed on the screened protein data to deal with missing values. Subsequently, Lasso regression method was used to determine the optimal regularization parameter *λ* through 10-fold cross-validation to further screen the combination of protein markers with the most predictive value, effectively reducing the data dimensionality and eliminating redundant variables.

In the data preprocessing stage, the study used stratified random sampling to divide the dataset into the training set and the test set in the ratio of 6:4 to ensure a balanced distribution of baseline features between the two groups. The random over sampling examples (ROSE) algorithm was used for adaptive sampling in order to optimize the data distribution by synthesizing a few class samples and undersampling the majority class samples, and at the same time, strictly limit the sampling operation to be performed only in the training set to avoid information leakage.

Based on the processed training data, three machine learning based prediction models were constructed in parallel. The Random Forest^[11]^ model was set up with 300 decision trees, the number of node split features was taken as the square root of the total number of features, and the out-of-bag error was used for internal validation. The LightGBM^[12]^ and XGBoost^[13]^ models were applied with the histogram optimization algorithm and the weighted quartile sketching algorithm, respectively, with an early stopping mechanism (patience = 100 round) to prevent overfitting. The hyper-parameters were optimized through 5-fold cross-validation. For feature importance assessment, Random Forest used the Gini impurity metric, while LightGBM and XGBoost were evaluated based on split gain.

We developed two progressive prediction models. The baseline model (Model 1) incorporated traditional risk factors (age, sex, BMI, smoking status, alcohol consumption, education level, Townsend deprivation index, and physical activity), while the enhanced model (Model 2) additionally included protein biomarkers obtained through screening. Given the established clinical utility of NT-proBNP in HF assessment, we specifically evaluated an intermediate model (Model 1 + NT-proBNP) for HF prediction alongside the two primary models. All models were validated on a randomly selected 40% holdout test set that preserved the original data distribution. For Model 2 integrating protein biomarkers, we rigorously compared three algorithms (Random Forest, LightGBM, XGBoost) and selected the top-performer based on cross-validated area under the receiver operating characteristic (ROC) curve (AUC), whereas both Model 1 and its NT-proBNP-enhanced model uniformly employed Random Forest to isolate the effects of traditional risk factors. This dual strategy optimally balances biomarker exploration with methodological control.

Model discriminative performance was assessed through ROC curve analysis with AUC quantification. We systematically compared the predictive performance of the two models using DeLong’s test, first examining AUC differences between the Model 1 and optimized Model 2. For HF prediction specifically, we further analyzed the discriminatory power between the enhanced Model 1 supplemented with NT-proBNP and the optimized Model 2.

We evaluated the predictive performance of the optimal Model 2 at predefined risk thresholds (5%, 10%, and 20%) by calculating sensitivity, specificity, positive predictive value (PPV), and negative predictive value (NPV) in the test data set. All metrics were derived based on whether the predicted probabilities exceeded the respective risk thresholds. To ensure reproducibility, analyses were conducted using a fixed random seed (seed = 123).

### Potential biomarker identification

For marker selection, the study employed a dual-verification approach for biomarker screening. First, protein biomarkers showing significant predictive performance were identified using the prediction model with the highest AUC. In parallel, proteins demonstrating independent prognostic associations were selected through multivariable Cox proportional hazards models (*P* < 1.71◊10^-5^). Biomarkers fulfilling both key criteria were ultimately chosen as candidate therapeutic targets.

All statistical analyses were performed using R software (version 4.4.1)^[14]^. Key analyses were conducted with the support of R packages including survival^[15]^, clusterProfiler^[16]^, rms^[17]^, cmprsk^[18]^, glmnet^[19]^, randomForest^[20]^, xgboost^[21]^ and lightgbm^[22]^.

## Results

### Baseline characteristics of participants

Table 1 presents the baseline characteristics of the study population stratified by outcome status. In the MI→all-cause mortality cohort, individuals who died were more likely to be older, have a history of smoking, and report physical inactivity (*P* < 0.001). Those who developed HF were generally older and had higher BMI (*P* < 0.05) in MI→HF cohort. In contrast, no significant differences in baseline characteristics were observed between survivors of MI with and without IS (*P* > 0.05).

### Associations of proteins with different outcomes in survivors of MI

Cox regression analyses suggested that multiple proteins showed statistically significant associations across outcomes after adjusting for multiple tests (*P* < 1.71◊ 10^-5^, Supplementary Data 3-5). Specifically, 570 proteins were significantly associated with all-cause mortality, including nine protective proteins (*e.g.*, UMOD, NELL1) and 561 risk-associated proteins (*e.g.*, EDA2R, TNFRSF10B) (Figure 2A). For HF, 172 proteins showed significant associations, consisting of 171 risk proteins (*e.g.*, NT-proBNP, NPPB) and one protective protein (UMOD) (Figure 2B). For IS, 13 associated proteins were identified, among which NT-proBNP and TIMP were significantly and positively associated with increased risk (Figure 2C).

**Figure 2.**
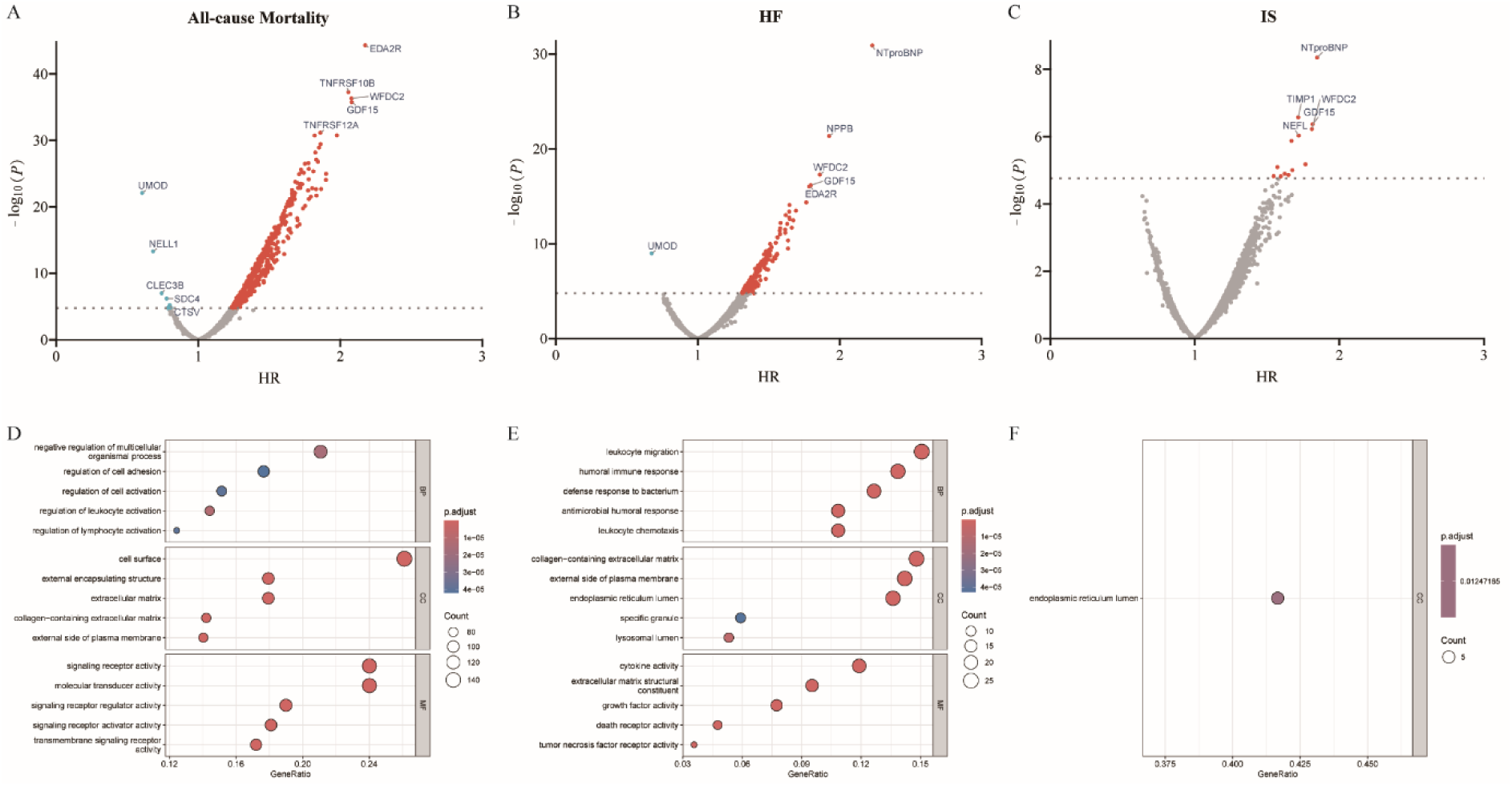
Associated proteins and the related enriched pathways. Volcano Plots for the correlated proteins for all-cause mortality (A), incident HF (B) and incident IS (C) in survivors of MI. The dash lines in A-C indicate the significance threshold of *P* 1.71×10^−5^. Grey dots indicate non-significant proteins, blue dots indicate significant protective proteins (HR < 1) and red dots indicates significant risk proteins (HR > 1). Top 5 significant risk and protective proteins are labeled. Pathway enrichment analysis results for the associated proteins for all-cause mortality (D), incident HF (E) and incident IS (F) in survivors of MI. BP: Biological Process; CC: Cellular Component; MF: Molecular Function. MI: Myocardial Infarction; HF: Heart Failure; IS: Ischemic Stroke.

For mortality-associated proteins, significant enrichment was observed in biological processes related to immune response (*e.g.*, positive regulation of cell adhesion, leukocyte migration), cellular components (*e.g.*, collagen-containing extracellular matrix, external side of plasma membrane), and molecular functions (*e.g.*, glycosaminoglycan binding, cytokine activity) (Figure 2D). Proteins associated with HF were primarily enriched in immune and antimicrobial-related processes, such as defense response to bacterium and antimicrobial humoral response, organelle-related components including the endoplasmic reticulum and extracellular matrix, and molecular functions like glycosaminoglycan binding and cytokine activity (Figure 2E). For IS-associated proteins, enrichment was mainly observed in biological processes related to inhibition of protease activity, including negative regulation of peptidase activity and negative regulation of hydrolase activity, cellular components such as the endoplasmic reticulum lumen and specific granule membrane, and molecular functions including endopeptidase inhibitor activity and peptidase inhibitor activity (Figure 2F). We also found 63 proteins showed significant nonlinear relationships (*P* < 1.71◊10^-5^), with 42 (66.7%) also demonstrating linear trends for all-cause mortality (Supplementary Data 6). Similarly, 22 proteins exhibited nonlinear associations with HF, half of which concurrently showed linear effects (Supplementary Data 7). However, no proteins met the significance threshold for nonlinear associations with IS. For all-cause mortality, AOC3 exhibited a J-shaped association (Figure 3A), suggesting a threshold concentration beyond which mortality risk rises sharply. IL12B displayed a U-shaped association (Figure 3B), indicating that both abnormally low and high concentrations may elevate via distinct pathological pathways. For HF, NT-proBNP showed a J-shaped association (Figure 3C), highlighting the potential value of its concentration threshold for HF risk stratification. ADAM12 had a U-shaped association with HF risk (Figure 3D), suggesting a possible role in HF pathogenesis through multiple mechanisms.

**Figure 3.**
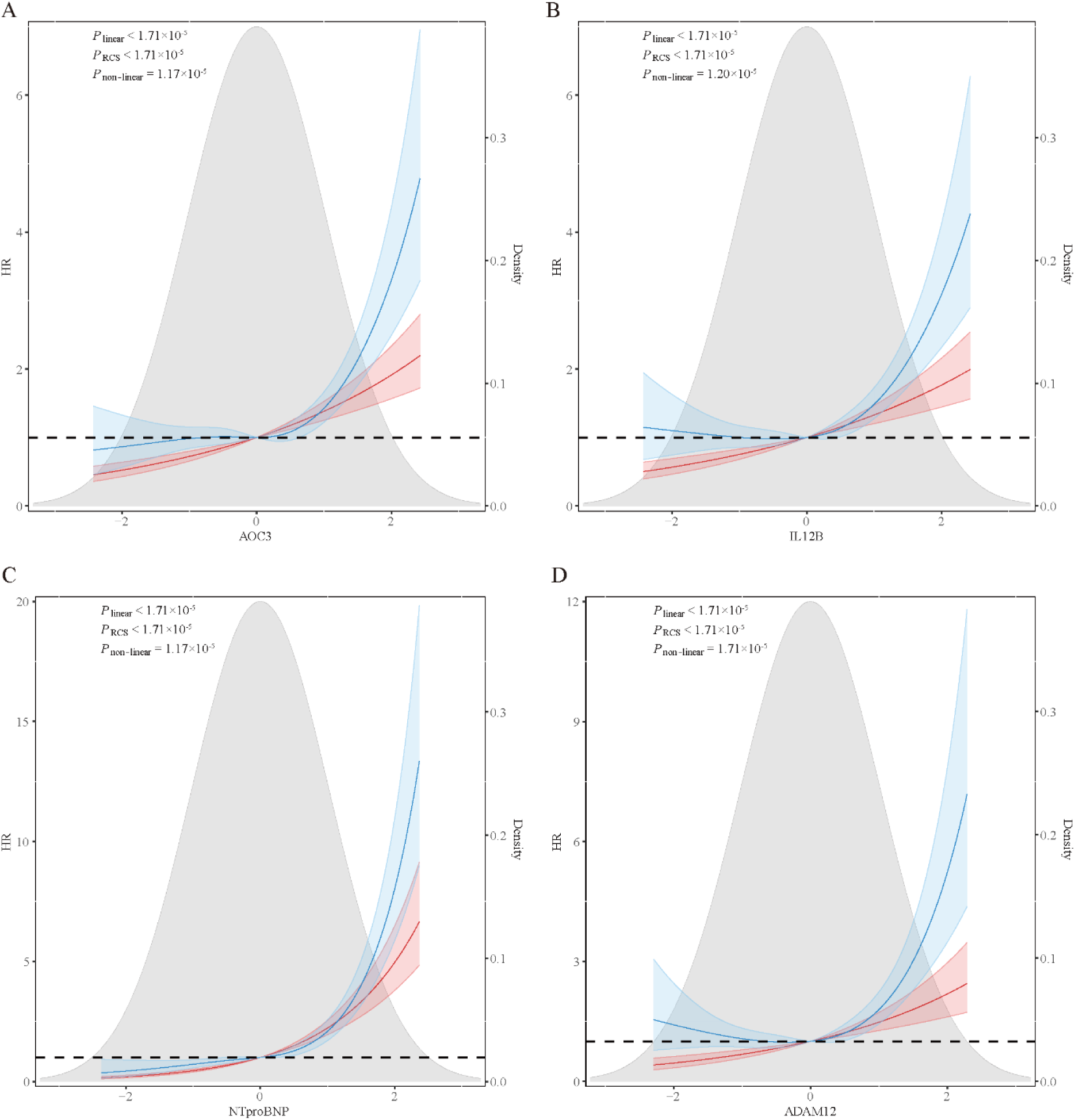
Restricted cubic spline analysis. This figure illustrates the nonlinear relationship between protein concentration and the risk of all-cause mortality (A, B) and HF (C, D). The J-shaped curves (A, C) showed that there was a concentration threshold effect, and the risk increased sharply after the threshold was exceeded. U-shaped curves (B, D) indicate an increased risk at both low and high concentrations. The red curve represents the linear Cox model results, and the blue curve shows the nonlinear RCS fit results (the shaded areas are 95% confidence intervals). The dotted line represents the reference line with HR of 1 and the gray background shows the protein concentration distribution density plot. The significance threshold was set to *P* < 1.71×10^−5^. HF: Heart Failure.

### Sensitivity analysis of protein-outcome associations

In the sex-stratified analysis, effect sizes were generally consistent between males and females, with Pearson correlation coefficients being 0.77 for all-cause mortality, 0.60 for HF, and 0.47 for IS (Figure 4A-4C). In the age-stratified analysis, age-related heterogeneity was small for all-cause mortality and HF, while medium heterogeneity was observed for IS, with Pearson correlation coefficient ranging from 0.22 for IS to 0.83 for all-cause mortality (Figure 4D-4F). Detailed hazard HR estimates for sex and age subgroup analyses are provided in Supplementary Figures 1-6.

**Figure 4.**
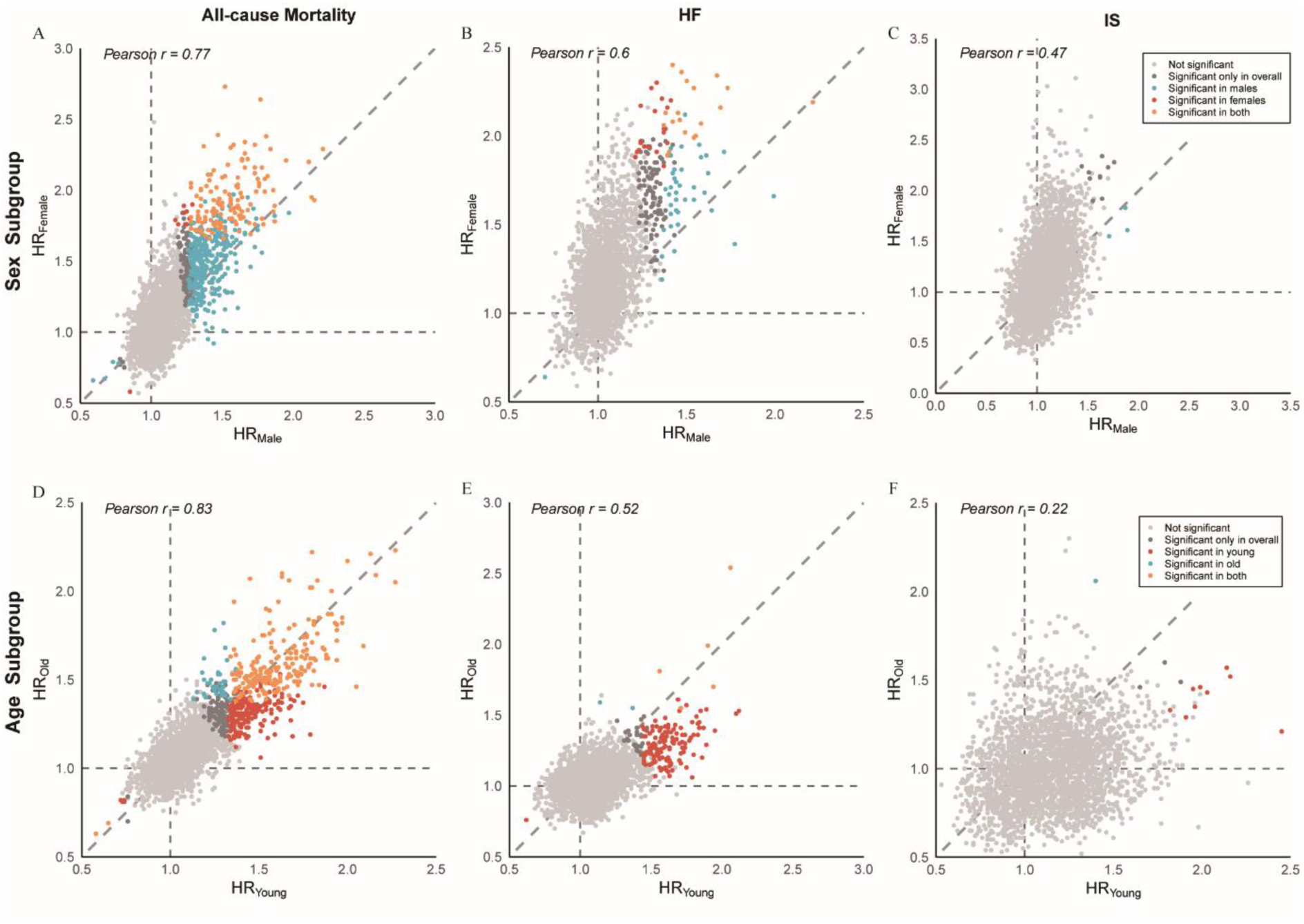
Subgroup analysis. Scatter plots comparing HR values between males (x-axis) and females (y-axis) for all-cause mortality (A), incident HF (B), and incident IS (C); and between young (<65 years, x-axis) and old (≥65 years, y-axis) patients for all-cause mortality (D), incident HF (E), and incident IS (F). Dashed lines indicate equivalent HRs. Pearson r is annotated top-left. Light gray dots indicate non-significant associations; dark gray dots represent proteins significant only in overall analysis; blue dots show associations significant in males/old patients only; red dots denote associations significant in females/young patients only; and orange dots indicate significance in both groups. HF: Heart Failure; IS: Ischemic Stroke.

Furthermore, to account for competing events in the MI cohort, Fine-Gray subdistribution hazard models were applied to assess the associations between protein levels and the risks of HF and IS, with death treated as a competing risk. The models were adjusted for the same covariates used in the linear Cox regression. Supplementary Figures 7-9 showed the cumulative incidence functions and suggested the HR agreement between Cox regression and competing risk models for HF and IS

### Proteomic risk prediction model for outcomes of MI survivors

Based on the nominal significant proteins (*P* < 0.05) in the initial screening of Cox regression, Lasso regression finally selected 117, 73 and 82 key proteins for constructing the prediction model for all-cause mortality, HF and IS, respectively. Comparative analysis of machine learning algorithms revealed differential predictive performance for distinct cardiovascular outcomes (Figure 5). For all-cause mortality prediction, the XGBoost algorithm demonstrated optimal discriminative ability (AUC = 0.79), while the LightGBM model showed superior predictive accuracy for HF (AUC = 0.81). Notably, the simplified model for HF incorporating only NT-proBNP showed relatively limited predictive efficacy (AUC = 0.71). The Random Forest algorithm exhibited the best performance in predicting IS (AUC = 0.76). DeLong’s test demonstrated that the proteomics-enhanced Model 2 achieved statistically and clinically superior discriminative performance compared to both Model 1 (ΔAUC: 0.13-0.21; *P* < 0.001) and Model 1 supplemented with NT-proBNP specifically for HF (ΔAUC: 0.10; *P* < 0.001). The machine learning based models demonstrating maximal AUC performance during internal validation were further evaluated in the test data set. Table 2 presents their comparative predictive performance at 5%, 10% and 20% risk thresholds with detailed sensitivity, specificity, positive predictive value, and negative predictive value results.

**Figure 5.**
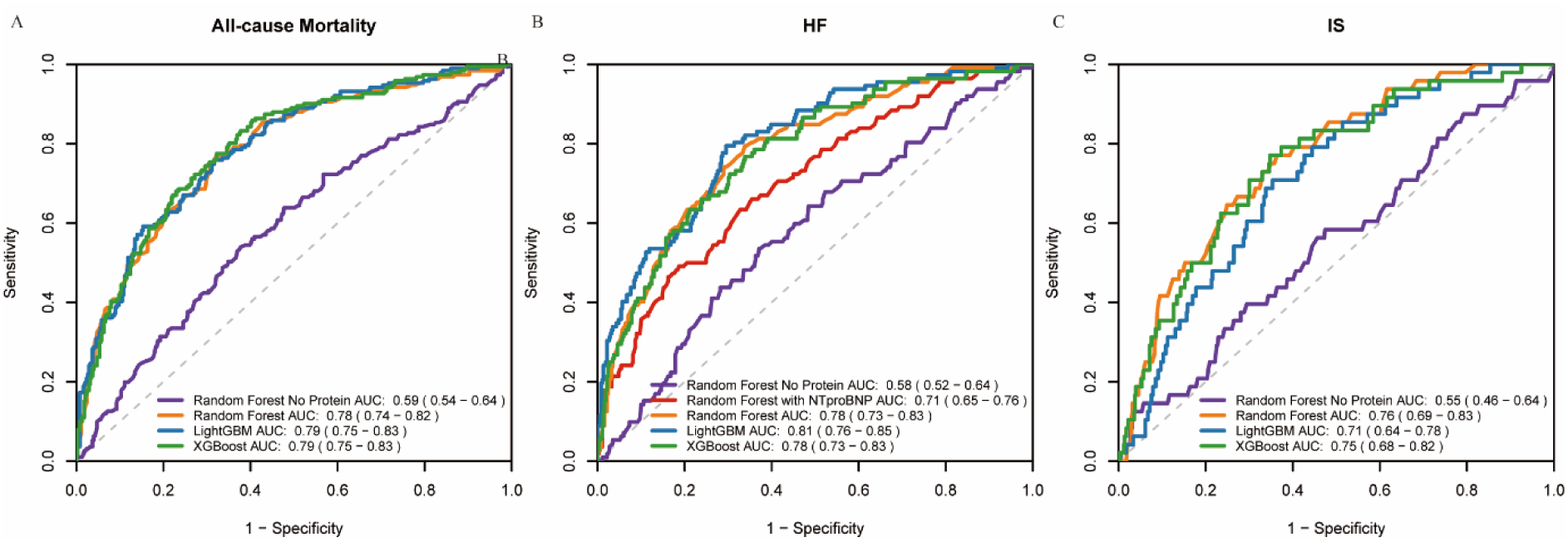
Performance evaluation for the machine learning-based models. ROC curves comparing predictive performance for all-cause mortality (A), incident HF (B) and incident IS (C) in the test set. Dashed diagonal lines represent reference (AUC = 0.5). Model performance is shown by: purple for Random Forest (model 1 without protein), orange for Random Forest (Model 2 with protein-enhanced), blue for LightGBM (Model 2 with protein-enhanced), green for XGBoost (Model 2 with protein-enhanced). ROC: Receiver operating characteristic; MI: Myocardial Infarction; HF: Heart Failure; IS: Ischemic Stroke; AUC: Area Under Curve.

**Table 2.** Performance of machine learning model for predicting clinical outcomes for survivors of MI at different predefined risk thresholds.

| Risk threshold, %<sup>a</sup> | No. of participants |  |  |  | Performance metric (95% CI) |  |  |  |
| --- | --- | --- | --- | --- | --- | --- | --- | --- |
|  | True-positive | False-negative | False-positive | True-negative | Sensitivity<sup>b</sup> | Specificity<sup>b</sup> | PPV<sup>b</sup> | NPV<sup>b</sup> |
| All-cause mortality (XGBoost AUC = 0.79) |  |  |  |  |  |  |  |  |
| ≥5 | 170 | 21 | 188 | 191 | 0.89 (0.84-0.93) | 0.50 (0.45-0.56) | 0.47 (0.42-0.53) | 0.90 (0.85-0.94) |
| ≥10 | 161 | 30 | 150 | 229 | 0.84 (0.78-0.89) | 0.60 (0.55-0.65) | 0.52 (0.46-0.57) | 0.88 (0.84-0.92) |
| ≥20 | 143 | 48 | 116 | 263 | 0.75 (0.68-0.81) | 0.69 (0.64-0.74) | 0.55 (0.49-0.61) | 0.85 (0.80-0.88) |
| HF (LightGBM AUC = 0.81) |  |  |  |  |  |  |  |  |
| ≥5 | 86 | 26 | 105 | 264 | 0.77 (0.68-0.84) | 0.72 (0.67-0.76) | 0.45 (0.38-0.52) | 0.91 (0.87-0.94) |
| ≥10 | 73 | 39 | 89 | 280 | 0.65 (0.56-0.74) | 0.76 (0.71-0.80) | 0.45 (0.37-0.53) | 0.88 (0.84-0.91) |
| ≥20 | 66 | 46 | 79 | 290 | 0.59 (0.49-0.68) | 0.79 (0.74-0.83) | 0.46 (0.37-0.54) | 0.86 (0.82-0.90) |
| IS (Random Forest AUC = 0.76) |  |  |  |  |  |  |  |  |
| ≥5 | 48 | 0 | 501 | 5 | 1.00 (0.93-1.00) | 0.01 (0.00-0.02) | 0.09 (0.07-0.11) | 1.00 (0.48-1.00) |
| ≥10 | 48 | 0 | 441 | 65 | 1.00 (0.93-1.00) | 0.13 (0.10-0.16) | 0.10 (0.07-0.13) | 1.00 (0.94-1.00) |
| ≥20 | 37 | 11 | 199 | 307 | 0.77 (0.63-0.88) | 0.61 (0.56-0.65) | 0.16 (0.11-0.21) | 0.97 (0.94-0.98) |

Note: Models were selected based on maximal AUC in internal validation. a. Risk thresholds (5%, 10%, 20%) represent the minimum predicted probability at which a participant is classified as high-risk. b. Sensitivity: Proportion of true events correctly identified; Specificity: Proportion of non-events correctly excluded; PPV: Probability that screen-positive individuals truly develop the outcome; NPV: Probability that screen-negative individuals remain outcome-free. MI: Myocardial Infarction; HF: Heart Failure; IS: Ischemic Stroke; CI: Confidence Interval; PPV: Positive Predictive Value; NPV: Negative Predictive Value.

The results of the feature importance analysis, as shown in Supplementary Figures 10-12, demonstrated a high degree of concordance between the top-ranking protein predictors identified by the machine learning models and those highlighted in the prior Cox regression analysis. We identified 52, 14 and four high-confidence protein markers from the intersection of significant proteins from Cox regression analyses and machine learning-selected features for all-cause mortality, HF, and IS, respectively. Specifically, TNFRSF10B and UMOD had the highest contribution to the prediction of all-cause mortality, NT-proBNP was the top predictive marker for HF, and also showed significant value in IS prediction. This cross-methodological validation not only reinforces the robustness of the findings, but also provides strong support for the biological relevance of these protein markers in post-MI risk stratification.

## Discussion

In this study, we identified a number of proteins significantly associated with major adverse clinical outcomes, including all-cause mortality, HF, and IS, in survivors of MI. Building upon these findings, we have developed machine learning based prediction models incorporating proteomic data, and further identified the potential prognostic protein markers.

For all-cause mortality, we identified 52 high-confidence protein markers. Notably, proteins involved in vascular homeostasis, such as ADA2, EDN1, and VEGFD, showed prominent dysregulation. ADA2, the predominant adenosine deaminase isoform in human plasma secreted by monocytes and macrophages, has been implicated in adverse clinical conditions including stroke, systemic vasculitis, immunodeficiency, and hematologic disorders^[23–25]^. Overexpression of endothelin-1 (ET-1), encoded by the EDN1 gene, a potent vasoconstrictor, is associated with coronary artery spasm, impaired myocardial perfusion, and progression of cardiovascular disease^[26,27]^, highlighting the pathological relevance of EDN1 in vascular dysfunction. MMP12, another key protein, contributes to vascular remodeling and atherosclerosis. Its overexpression has been linked to chronic diseases such as chronic obstructive pulmonary disease (COPD) and emphysema^[28]^, however, its specific role in post-MI mortality warrants further investigation. In addition, the identification of renal-cardiac interaction markers, such as UMOD and REN, underscores the contribution of renal dysfunction to cardiovascular prognosis. UMOD influences volume regulation via renal sodium handling^[29]^, while REN, reflecting RAAS overactivation, is implicated in hypertension, HF, and metabolic dysregulation. These findings support the concept of cardiorenal-metabolic (CKM) syndrome, where renal, cardiovascular, and metabolic pathways interact, providing a molecular basis for integrated risk assessment and management^[30]^. Finally, the enrichment of proteins associated with inflammation and apoptosis, including TNFRSF10B, FCN1, and LILRB2, suggests that dysregulation of innate immunity and programmed cell death may contribute to multi-organ failure in the post-MI setting.

For outcome of HF in survivors of MI, we identified 14 core protein markers. Among these, NT-proBNP, a gold-standard biomarker for ventricular wall stress assessment, demonstrated robust predictive performance in both Cox proportional hazards analyses and machine learning frameworks, corroborating its well-documented clinical utility. A particularly novel finding was the identification of IGFBP7, a protein that modulates insulin-like growth factor (IGF) bioavailability and has been implicated in HF with preserved ejection fraction (HFpEF)^[31]^. The overexpression of CD74, a chaperone for major histocompatibility complex class II molecules, points to the involvement of antigen-presenting cells in myocardial inflammation^[32]^. This observation provides a theoretical basis for investigating immunomodulatory strategies, such as targeting macrophage polarization. In addition, the detection of GDF-15 and IL-6 further emphasizes the central role of inflammation and cellular stress in HF progression. GDF-15, a member of the TGF-β cytokine superfamily, is involved in myocardial remodeling and has been shown to be elevated in both acute myocardial infarction and chronic HF^[33,34]^. IL-6, meanwhile, is considered a key inflammatory mediator in HFpEF, contributing to systemic inflammation and diastolic dysfunction^[35]^.

In the context of IS prediction, four highly specific protein markers, offering novel insights into heart-brain interactions. The re-identification of NT-proBNP reinforces the role of cardiac dysfunction as a key risk factor for embolic stroke, potentially mediated by blood stasis and thrombus formation due to impaired left ventricular systolic function. OLR1, a scavenger receptor for oxidized low-density lipoproteins, plays a dual role in the pathogenesis of atherosclerosis. It not only contributed to plaque formation, but also promoted plaque instability through the induction of foam cell accumulation^[36]^. Elevated levels of NEFL, a neuron-specific intermediate filament protein, in peripheral blood may indicate subclinical cerebral small-vessel disease, which increases susceptibility to ischemic events^[37]^. NPPB is the precursor of brain natriuretic peptide, and the concurrent detection of NPPB alongside NT-proBNP suggests that full activation of the neurohumoral system may serve as a common pathogenic pathway underlying cardiac-cerebral comorbidities.

We revealed significant nonlinear associations between certain serum protein levels and disease risk through restricted cubic spline analysis. To further explore this complex dose-response relationship, we employed machine learning-based regression algorithms, which better captured complex interactions among variables and potential nonlinear patterns. This approach provided a more precise quantitative basis for elucidating the biological mechanisms of serum proteins in disease development and progression. Furthermore, we demonstrated that integrating protein biomarkers with conventional risk factors enhanced risk stratification for all-cause mortality, HF, andIS in myocardial infarction survivors. The optimized model exhibited threshold-dependent performance: superior sensitivity at the 5% threshold enabled early detection of high-risk individuals, while enhanced positive predictive value at the 20% threshold better identified candidates for resource-intensive interventions. These differential performance characteristics underscore the necessity of context-specific implementation in clinical practice.

This study has several limitations that warrant consideration. First, the study population comprised only European descendants, and the generalizability of the findings to other ethnic groups requires further validation. Second, endpoint events were primarily defined based on registry data, which may introduce some degree of misclassification. Third, variations in the time intervals between blood sample collection and protein quantification may introduce preanalytical variability, although all samples underwent standardized processing protocols. Notably, restricted cubic spline analyses revealed nonlinear associations for certain proteins, suggesting that threshold effects might be overlooked by linear models. Subgroup analyses revealed lower heterogeneity in protein-outcome associations for all-cause mortality and HF populations, but higher heterogeneity in IS cases, potentially due to the limited sample size of IS participants. Furthermore, competing risk analyses showed that some proteins significant in Cox models lost statistical significance after accounting for competing events. Nevertheless, the multimodal approach integrating machine learning with traditional survival analysis demonstrated good robustness in this study.

## Conclusion

This study systematically identified 52 mortality-associated, 14 HF-associated, and 4 IS-associated plasma protein biomarkers in survivors of MI. These findings offer valuable insights into the biological mechanisms underlying post-MI prognosis and support the potential utility of proteomic markers in future risk stratification and personalized management strategies.

## Funding

This work is supported by National Natural Science Foundation of China (No. 32470658) and the Fundamental Research Funds for the Central Universities.

## Data Availability

The data analyzed in this study were obtained from the UK Biobank (application number: 88159). The research data are available through application to the UK Biobank (https://www.ukbiobank.ac.uk/). The authors would like to express their sincere gratitude to all UK Biobank participants and the staff who contributed to this research.

## Acknowledgments

We gratefully thank to all the participants involved in the UK Biobank. We thank all staff at UK Biobank for their contributions to this research.

## Conflict of interest

The authors declare they have no conflict of interest.

## Author contributions

Conceptualization: Xingjie Hao, Bofang Zhang, Shizhen Xiang, Yuge Ye

Methodology: Xingjie Hao, Shizhen Xiang, Yuge Ye, Xi Cao, Huidan Zeng

Investigation: Shizhen Xiang, Yuge Ye, Yunlong Guan, Siyu Zhu

Visualization: Shizhen Xiang, Yuge Ye, Xingjie Hao

Funding acquisition: Xingjie Hao, Bofang Zhang

Project administration: Xingjie Hao, Bofang Zhang

Supervision: Xingjie Hao, Bofang Zhang

Writing – original draft: Shizhen Xiang, Yuge Ye

Writing – review & editing: Shizhen Xiang,Yuge Ye, Xingjie Hao, Bofang Zhang, Xi

Cao, Huidan Zeng, Yunlong Guan, Siyu Zhu, Xiangjing Liu, Da Luo, Yifan Kong,

Zhonghe Shao

## Notes

### Competing Interest Statement

The authors have declared no competing interest.

### Clinical Trial

This study is not applicable.

### Author Declarations

All participants provided written informed consent, and the study was approved by the North West Multi-Center Research Ethics Committee (Research Ethics Committee reference: 11/NW/0382).

## References

[1] Yeh RW, Sidney S, Chandra M, Sorel M, Selby JV, Go AS. Population trends in the incidence and outcomes of acute myocardial infarction. N Engl J Med. 2010;362(23):2155–2165.

[2] Hahla M S, Saeed Y, Razieh H. Comparison of risk factors & clinical and angiographic characterization of STEMI in young adults with older patients[J]. Research journal of pharmaceutical biological and chemical sciences, 2016, 7(6): 2013–2016.

[3] Saito Y, Oyama K, Tsujita K, Yasuda S, Kobayashi Y. Treatment strategies of acute myocardial infarction: updates on revascularization, pharmacological therapy, and beyond. J Cardiol. 2023;81(2):168–178.

[4] Lu B, Posner D, Vassy JL, et al. Prediction of Cardiovascular and All-Cause Mortality After Myocardial Infarction in US Veterans. Am J Cardiol. 2022;169:10–17.

[5] Takeuchi S, Honda S, Nishihira K, et al. Prognostic impact of heart failure admission in survivors of acute myocardial infarction. ESC Heart Fail. 2024;11(4):2344–2353.

[6] Chan MY, Efthymios M, Tan SH, et al. Prioritizing Candidates of Post-Myocardial Infarction Heart Failure Using Plasma Proteomics and Single-Cell Transcriptomics [published correction appears in Circulation. 2020 Oct 13;142(15): e234.

[7] Castiglione V, Aimo A, Vergaro G, Saccaro L, Passino C, Emdin M. Biomarkers for the diagnosis and management of heart failure.Heart Fail Rev. 2022;27(2):625–643.

[8] Monbailliu T, Goossens J, Hachimi-Idrissi S. Blood protein biomarkers as diagnostic tool for ischemic stroke: a systematic review. Biomark Med. 2017;11(6):503–512.

[9] Sun Z, Yun Z, Lin J, et al. Comprehensive mendelian randomization analysis of plasma proteomics to identify new therapeutic targets for the treatment of coronary heart disease and myocardial infarction. J Transl Med. 2024;22(1):404. Published 2024 Apr 30.

[10] Qin C, Zhao XL, Ma XT, et al. Proteomic profiling of plasma biomarkers in acute ischemic stroke due to large vessel occlusion. J Transl Med. 2019;17(1):214. Published 2019 Jul 1.

[11] Breiman, L. “Random Forests.” Machine Learning 45 (2001): 5–32.

[12] Ke, Guolin, et al. “LightGBM: A Highly Efficient Gradient Boosting Decision Tree.” Neural Information Processing Systems (2017).

[13] Chen, Tianqi, and C. Guestrin. “XGBoost: A Scalable Tree Boosting System.” ACM (2016).

14. [14] R Core Team (2024). _R: A Language and Environment for Statistical Computing_. R Foundation for Statistical Computing, Vienna, Austria. <https://www.R-project.org/>.

[15] Therneau T (2024). A Package for Survival Analysis in R. R package version 3.8-3. <https://CRAN.R-project.org/package=survival>.

[16] Wu T, Hu E, Xu S, Chen M, Guo P, Dai Z, Feng T, Zhou L, Tang W, Zhan L, Fu X, Liu S, Bo X, Yu G. clusterProfiler 4.0: A universal enrichment tool for interpreting omics data. Innovation (Camb). 2021 Jul 1;2(3):100141. doi: 10.1016/j.xinn.2021.100141. PMID: 34557778; PMCID: PMC8454663.

[17] Harrell FE Jr (2024). rms: Regression Modeling Strategies. R package version 7.1-1. <https://CRAN.R-project.org/package=rms>.

[18] Gray B (2024). cmprsk: Subdistribution Analysis of Competing Risks. R package version 2.2-11. <https://CRAN.R-project.org/package=cmprsk>.

[19] Friedman J, Hastie T, Tibshirani R. Regularization Paths for Generalized Linear Models via Coordinate Descent. J Stat Softw. 2010;33(1):1–22. PMID: 20808728; PMCID: PMC2929880.

[20] Liaw A, Wiener M (2002). “Classification and Regression by randomForest.” R News, 2(3):18–22. <https://CRAN.R-project.org/package=randomForest>.

[21] Chen T, Guestrin C (2016). “XGBoost: A Scalable Tree Boosting System.” In Proceedings of the 22nd ACM SIGKDD International Conference on Knowledge Discovery and Data Mining, 785–794.

[22] Ke G, Meng Q, Finley T, Wang T, Chen W, Ma W, Ye Q, Liu T-Y (2017). “LightGBM: A Highly Efficient Gradient Boosting Decision Tree.” In Advances in Neural Information Processing Systems, 30. <https://github.com/microsoft/LightGBM>.

[23] Wu Z, Gao S, Watanabe N, et al. Single-cell profiling of T lymphocytes in deficiency of adenosine deaminase 2. J Leukoc Biol. 2022;111(2):301–312.

[24] Lee PY, Schulert GS, Canna SW, et al. Adenosine deaminase 2 as a biomarker of macrophage activation syndrome in systemic juvenile idiopathic arthritis. Ann Rheum Dis. 2020;79(2):225–231.

[25] Pinto B, Deo P, Sharma S, Syal A, Sharma A. Expanding spectrum of DADA2: a review of phenotypes, genetics, pathogenesis and treatment. Clin Rheumatol. 2021;40(10):3883–3896.

[26] Halcox JP, Nour KR, Zalos G, Quyyumi AA. Endogenous endothelin in human coronary vascular function: differential contribution of endothelin receptor types A and B. Hypertension. 2007;49(5):1134–1141.

[27] Lanza GA, Crea F. Primary coronary microvascular dysfunction: clinical presentation, pathophysiology, and management. Circulation. 2010;121(21):2317–2325.

[28] Gharib SA, Manicone AM, Parks WC. Matrix metalloproteinases in emphysema. Matrix Biol. 2018;73:34–51.

[29] Wuttke M, Li Y, Li M, et al. A catalog of genetic loci associated with kidney function from analyses of a million individuals. Nat Genet. 2019;51(6):957–972.

[30] Sebastian SA, Padda I, Johal G. Cardiovascular-Kidney-Metabolic (CKM) syndrome: A state-of-the-art review. Curr Probl Cardiol. 2024;49(2):102344.

[31] Insulin-Like Growth Factor-Binding Protein-7 as a Biomarker of Diastolic Dysfunction and Functional Capacity in Heart Failure With Preserved Ejection Fraction: Results From the RELAX Trial. JACC Heart Fail. 2016;4(11):860–869.

[32] Su H, Na N, Zhang X, Zhao Y. The biological function and significance of CD74 in immune diseases. Inflamm Res. 2017;66(3):209–216.

[33] Wollert KC, Kempf T, Peter T, et al. Prognostic value of growth-differentiation factor-15 in patients with non-ST-elevation acute coronary syndrome. Circulation. 2007;115(8):962–971.

[34] Kempf T, Björklund E, Olofsson S, et al. Growth-differentiation factor-15 improves risk stratification in ST-segment elevation myocardial infarction. Eur Heart J. 2007;28(23):2858–2865.

[35] Alogna A, Koepp KE, Sabbah M, et al. Interleukin-6 in Patients With Heart Failure and Preserved Ejection Fraction. JACC Heart Fail. 2023;11(11):1549–1561.

[36] Tian K, Ogura S, Little PJ, Xu SW, Sawamura T. Targeting LOX-1 in atherosclerosis and vasculopathy: current knowledge and future perspectives. Ann N Y Acad Sci. 2019;1443(1):34–53.

[37] Peters N. Neurofilament Light Chain as a Biomarker in Cerebral Small-Vessel Disease. Mol Diagn Ther. 2022;26(1):1–6.

